# Metabolite-specific Reproducibility of Cerebral ^31^P-MRS at 3T: Recommendations for Clinical Research

**DOI:** 10.64898/2026.01.15.26343920

**Authors:** Magnus Svensen, Christian Dölle, Brage Brakedal, Haakon Berven, Njål Brekke, Alexander Richard Craig-Craven, Erika Veslemøy Sheard, Arve Hjellbrekke, Vivian Skjeie, John Georg Seland, Charalampos Tzoulis, Frank Riemer

## Abstract

Phosphorus magnetic resonance spectroscopy (^31^P-MRS) enables non-invasive measurement of brain metabolism, yet its reproducibility in clinical settings remains unclear. We systematically assessed intra- and intersession variability as well as inter-individual differences of key phosphorus metabolites at 3 Tesla in healthy individuals and persons with Parkinson’s disease under various experimental condition. Intersession variability, as measured by coefficients of variation (CoV) increased notably for longer scan intervals (∼1 year), and metabolite ratios from well-resolved spectral signals (i.e., adenosine triphosphate (ATP), phosphocreatine (PCr), intracellular inorganic phosphate P_i_) exhibited consistently higher stability compared to ratios calculated from metabolite signals overlapping on the spectrum (e.g., total nicotinamide adenine dinucleotide (tNAD), as well as phosphate monoesters (PMEs) and phosphate diesters (PDEs). Test-retest variability ranged from ∼5-25 CoV%, where PCr, ATP-α and ATP-γ were the most stable while glycerophosphocholine (GPC), glycerophosphoethanolamine (GPE), phosphoethanolamine (PE) and tNAD varied considerably. Inter-individual variability was found to be higher than intra-individual variability for all metabolite ratios, ranging from ∼9-33 CoV%. By systematically quantifying intra-individual and inter-individual variability, as well as providing explicit sample-size recommendations, this study facilitates more reliable longitudinal and cross-sectional clinical trials and translational studies of brain metabolism featuring ^31^P-MRS.

## Introduction

Disturbances in brain energy metabolism are central to the pathophysiology of numerous neurological disorders. Processes such as mitochondrial dysfunction, altered phospholipid turnover, and redox imbalance are tightly linked to altered cerebral bioenergetics^1–3^. Understanding these metabolic disturbances is therefore essential for elucidating disease mechanisms, monitoring progression, and evaluating therapeutic interventions. Phosphorus magnetic resonance spectroscopy (³¹P-MRS) is a non-invasive method that measures energy-related and membrane-related metabolites in the living human brain, enabling unique insights into brain aging and disease^4,5^. Key signals include adenosine triphosphate (ATP), phosphocreatine (PCr), inorganic phosphate (P_i_), phosphomonoesters (PME: phosphocholine [PC], phosphoethanolamine [PE]), phosphodiesters (PDE: glycerophosphocholine [GPC], glycerophosphoethanolamine [GPE]), and total nicotinamide adenine dinucleotide (tNAD, NAD[ and NADH)^6–8^. Other aggregated metabolite signals, such as ATP (sum of α, β, and γ resonances) and high-energy phosphates (HEP: sum of ATP, PCr, P_i_, tNAD), are also reported^4,9,10^.

Over the past decades, ³¹P-MRS has been used to study both rare monogenic disorders and common neurodegenerative diseases, including Alzheimer’s disease (AD) and Parkinson’s disease (PD)^4^. A recent meta-analysis revealed an elevated PME/PDE ratio has been reported in AD, whereas PD studies more often show reduced PME/PDE and PCr/Pi ratios^4^.

More recently, ³¹P-MRS has been incorporated into randomized clinical trials (RCTs) to assess therapeutic intervention. Examples include measuring brain metabolism indices as a proxy of mitochondrial function in a trial of ursodeoxycholic acid (UDCA) and assessing cerebral NAD levels in a trial of the NAD precursor nicotinamide riboside (NR)^11,12^. In both studies, ³¹P-MRS was central to demonstrating target engagement, guiding the decision to advance these therapies to larger trials. Another recent clinical trial employed ^31^P-MRS to demonstrate homeostatic effects on brain bioenergetics in persons with Parkinson’s disease (PwPs) following treatment with the nanocatalytic drug CNM-Au8^13^.

Despite its promise, ^31^P-MRS faces certain limitations that constrain its broader use in clinical research. It has lower sensitivity compared to the more commonly used ^1^H-MRS, largely due to the lower gyromagnetic ratio and lower *in vivo* concentrations of phosphorus compounds^5,14^. In addition, MRS methods are generally sensitive to long acquisition times and motion artifacts, which may increase variability in metabolite quantification^15^.

To enable meaningful application of ^31^P-MRS in observational and interventional studies, robust estimates of test-retest reproducibility and sample size requirements are needed. However, few reproducibility studies in the brain have been conducted, some of which report the reproducibility of only a single (or a few) metabolite signals and often feature single-session design. Reported coefficients of variation (CoVs) ranged from 1.2% to 32.6 % at 3T^9,16,17^. At 7 T, the CoVs for α-ATP, γ-ATP, PCr, P_i_, tNAD, PC, PE, GPC and GPE were reported to range from 4.4 % to 17.8 %^13,18^.

Previously reported ^31^P-MRS findings have also be contradictory^4,5,19^, for example, with respect to age-related changes in ATP^2,20–22^. This possibly reflects methodological heterogeneity in acquisition, pre-processing, quantification (absolute vs. ratio-based), quality control, small sample sizes, as well as the use of ATP as an internal concentration reference for other metabolites, with an assumed brain concentration of ∼3 mM^23^. However, since ATP concentrations may themselves be altered in disease^24–26^, this practice introduces further uncertainty into metabolite assessments. While some methodological considerations have been addressed in the literature^5^, more quantitative guidance on reproducibility and sample size requirements for ^31^P-MRS studies is needed.

To address this gap, we conducted a systematic assessment of between- and intra-individual reproducibility in cerebral ^31^P-MRS using four independent datasets acquired in a 3T scanner. These include: (1) repeat acquisitions without repositioning; (2) repeat scans with repositioning within 24 hours; (3) longitudinal scans of healthy volunteers over 49 weeks; and (4) test-retest scans from persons with Parkinson’s disease (PwPs) followed over 4 weeks. We quantified the variability of both well-resolved and overlapping metabolite ratios, assessed voxel placement consistency, and derived sample size recommendations for clinical and interventional research applications. Together, these results provide practical benchmarks for robust study design and interpretation in future ^31^P-MRS research.

## Materials and Methods

### Datasets

Four ^31^P-MRS datasets were used in our analyses (Figure 1), selected to cover a wide range of scenarios, including disease status, effect of leaving the scanner between visits (thus requiring new pre-scan calibrations, coil positionings and region of interest (ROI) placements) and having the same scan repeated, without re-positioning. All four datasets featured identical MRS parameters and were acquired on the same system.

**Figure 1.**
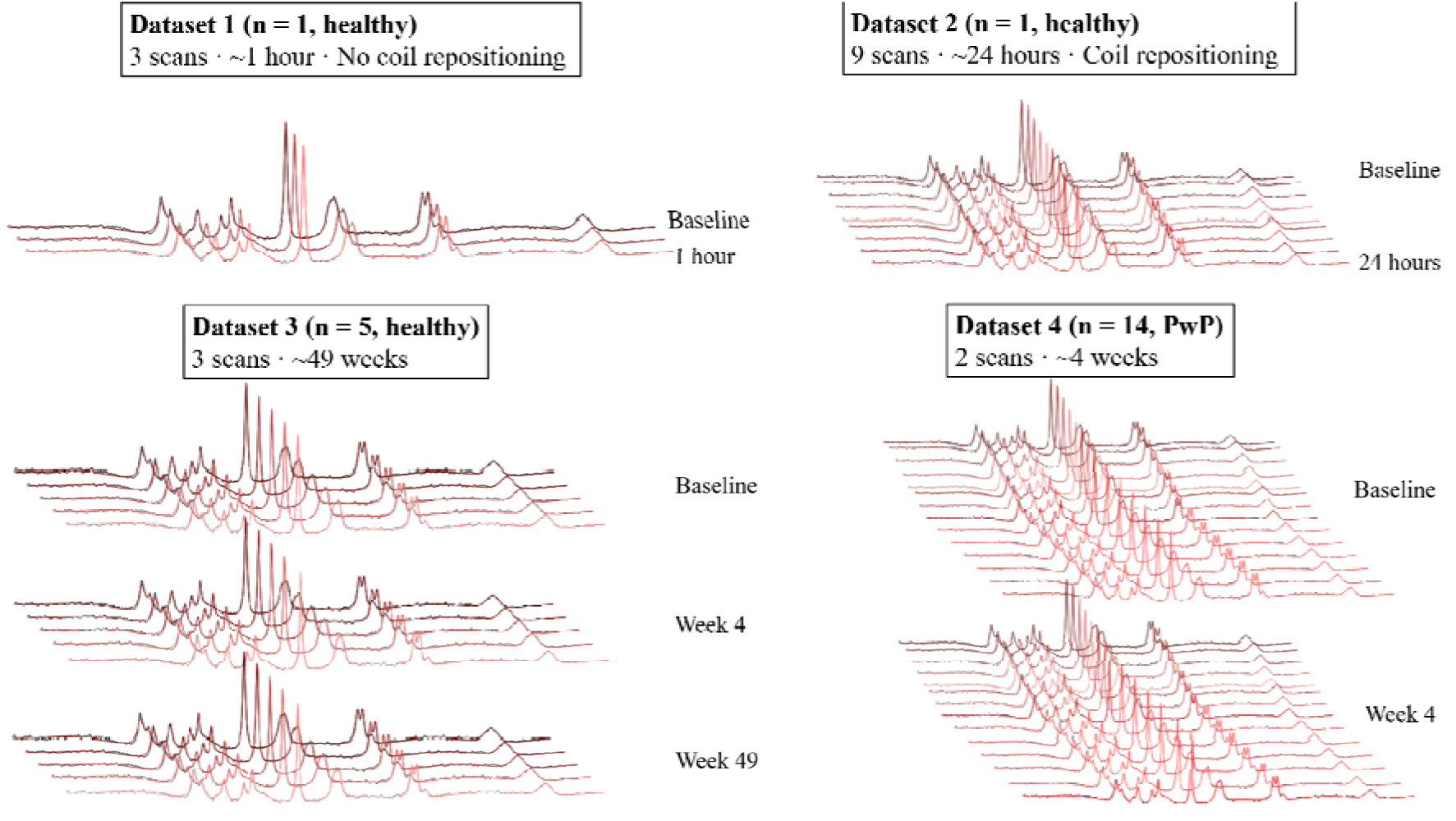
Schematic overview of data sets included in study. Data set one featured one neurologically healthy (control) male (n = 1) in their 40s who was scanned using ^31^P-MRS three times with no breaks or coil repositoning between the scans. Data set two featured one healthy male (n = 1) in their 40s scanned nine times over the course of 24 hours at timepoints 0, 0.5, 1, 2, 3, 4, 6, 8, and 23.5 hours. Data set three featured healthy participants (n = 5, 3M/2F, average age 44.6 + 1.5 years) scanned at baseline and approximately 4 weeks and 49 weeks later. Data set four featured participants with Parkinson’s disease (PwPs) (n = 14, 13M/1F, average age 63.7 ± 11.0 years) scanned at baseline and approximately 4 weeks later.

**Dataset one** featured a neurologically healthy male subject (n = 1) in their 40s scanned continuously for three acquisitions *without leaving the scanner.* The total scan time was approximately one hour.

**Dataset two** featured a neurologically healthy male subject (n = 1) in their 40s scanned with leaving the scanner and being re-positioned each time, for a total of nine timepoints over the course of approximately 24 hours at varying intervals (t = 0, 0.5, 1, 2, 3, 4, 6, 8, 23.5h).

**Dataset three** featured baseline data from a longitudinal study (the NAD-brain trial, ClinicalTrials.gov identifier: NCT05698771), in which healthy subjects (n = 5, 3M/2F, Age 44.6 ± 1.5 years) received three scans spread-out in time (at timepoints 0, approximately 4 weeks, and approximately 49 weeks).

**Dataset four** featured persons with Parkinson’s disease (PwPs) (n = 14, 13M/1F, Age 63.7 ± 11.0 years)) scanned at baseline and after approximately 1 month^12^.

### Ethics statement

All subjects provided written informed consent, and acquisition was in accordance with good clinical practice principles and the declaration of Helsinki. The collection of dataset one was approved by Haukeland University Hospital, Department of Radiology under protocol D70079 – MR examination of healthy volunteers for quality assurance. This protocol has ethical approval from the Regional Committees for Medical and Health Research Ethics Western Norway (REK Vest). The collection of datasets two, three and four was approved by the Regional Committees for Medical and Health Research Ethics Western Norway (REK Vest) with identifiers: REK Vest 496197 (NAD-brain study), REK Vest 2018/597 (NADPARK study). Both studies are registered on Clinicaltrials.gov with identifiers NCT05698771 and NCT03816020.

### Acquisition

All subjects were scanned with a 3 T Biograph mMR PET/MR scanner (Siemens Healthineers, Erlangen, Germany) equipped with a commercially available dual-tuned Tx/Rx ^1^H/^31^P birdcage coil (RAPID Biomedical, Rimpar, Germany).

An anatomical T_1_-weighted image with the following sequence parameters was acquired for positioning: MPRAGE 3D T_1_-weighted sagittal volume, TE/TR/TI = 2.26 ms /2400 ms /900 ms, acquisition matrix = 256 x 256 x 192, FOV = 256 x 256 x 192 mm^3^, 200 Hz/px readout bandwidth, flip angle = 8° and total acquisition duration of 5.6 m.

Whole-brain ^31^P-MRS was acquired using a vendor-provided 3D CSI FID product-sequence with the following sequence parameters: 8 x 8 matrix size, nominal voxel size 30 x 30 x 80 mm^3^, field of view (FOV) = 240 x 240 x 80 mm^3^, TE/TR = 2.3 ms / 3000 ms, 10 averages, non-selective RF excitation pulse flip angle = 90°, 1024 samples, readout length = 512 ms, 1000 Hz bandwidth and total acquisition time 14.5 m. For each subject, the 3D CSI FOV was centred on the brain midline and aligned parallel to the anterior and posterior commissure^12^.

Rectangular NOE pulses of 10 ms length, inter-pulse delay 1ms, train length 10 prior and WALTZ4 decoupling (2 ms pulses, 180 deg. flip angle) were applied prior to ^31^P-excitation and during the first half of the acquisition window respectively^12,27,28^.

### Spectral fitting and quantification

Spectral data were processed by a single fixed rater to eliminate interrater variability using the OXSA (*OXford S*pectral *A*nalysis) toolbox, which is a MATLAB-based, open-source toolbox for performing non-linear least squares fitting of MRS signals based on the AMARES algorithm^29,30^. Initial values for prior information were based upon literature values^6,27^. NAD^+^ and NADH were fitted separately and summed as tNAD for analysis. A complete overview of prior knowledge parameters is included as supplementary information (Appendix 2).

In order to correct for movement-related spectral shifts, as well as filtering voxels with poor SNR from the spectrum, a *post hoc* spectral registration filter was applied using a modified version of the function from the Gannet MRS toolbox^31,12^ to align the individual voxel spectra before quantification. Voxels in the occipital region of each participant were selected manually based on a combination of visual inspection of spectral quality and SNR. The Dice-Sørensen coefficient (DSC) was calculated to assess any differences in voxel selection between the scans. Areas of multiplet signals and grouped metabolites (PME, PDE, combined ATP, HEP) were summed. The individual ATP resonances (-α, -β, -γ) were normalized to PCr, while the other individual metabolite signals were normalized to ATP-α. In addition to calculating individual metabolite ratios, we also included the following metabolite ratios used in the meta-analysis by Jing *et al*.^4^: combined ATP/P_i_ (ATP = ATP-α, -β and -γ), HEP/P_i_(HEP = high energy phosphates, ATP-α, β, γ + PCr + tNAD), PCr/P_i_, PME/P_i_ (phosphate monoesters; PC and PE), PDE/P_i_ (phosphate diesters; GPC and GPE) and PME/PDE.

In our spectral analysis we decided to group metabolite signals based on the degree of which they overlap on the ^31^P-MR spectrum. Well-resolved metabolites comprise ATP-α,β,γ, PCr, P_i_ as well as aggregated signals from these measures (i.e., HEP and combined ATP). Overlapping metabolites comprise the rest of the spectrum; tNAD, GPC, GPE, PC, PE, MP and aggregate signals (i.e., PME and PDE).

### Statistical analysis

Normality was assessed by visual inspection of quantile–quantile (Q–Q) plots. Given the small sample size in our study, these were interpreted as descriptive diagnostics rather than formal decision criteria (Appendix 1). We evaluated the intra-individual variability within and between sessions using a combination of CoVs, intraclass correlation coefficients (ICC), Pearson’s correlations (*r*) and Bland-Altman plots^32,33^, where applicable. In this study, we defined a session as scans occurring within a short time frame. As such, *intrasession* variability (CoV) was assessed in datasets one and two, whereas *intersession* variability (CoV) was assessed in datasets three and four. In data sets three and four, individual intra-individual CoVs were calculated and then averaged. In line with a previous study by Pratt et al.^34^, we set the following thresholds for interpreting CoVs: < 10%, low; 10%-19%, moderate; 20%-29%, high; ≥30%, very high. For Pearson’s *r*: 0-0.9, no correlation; 0.1-0.39, weak correlation; 0.40-0.74, moderate correlation; 0.75-0.89, strong correlation; ≥ 0.9, very strong correlation. For ICCs: < 0.5 poor reliability; 0.5-0.74, moderate reliability; 0.75-0.89, good reliability; ≥0.9 very good reliability.

The intraclass correlation coefficients were calculated in MATLAB using the ICC(A-k) function (Arash Salarian, 2008). The ICC(A-k) measures absolute agreement for averaged measurements based on k independent measurements on randomly selected objects^35^. CoVs for studies one and two were calculated by taking the average and standard deviation for all measurements. In datasets three and four, individual intra-individual CoVs were calculated and averaged for each metabolite ratio in order to minimize the effect of group level baseline metabolite variations. Bland–Altman plots were constructed for each metabolite ratio of interest to assess the presence of systematic bias or proportional error between sessions.

We performed sample size calculations for (i) a two-tailed independent samples *t*-test using all of our data and (ii) a matched pairs *t*-test using pair-wise data from datasets 3 and 4 (baseline and week 4). Effect sizes and sample sizes were calculated in Excel and validated in Gpower^36^ assuming a significance level of α = 0.05 with varying percentage power, ranging from 50% to 95%.

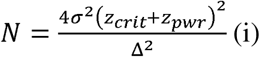

Where *N* is the total sample size (both groups combined); σ is the standard deviation of measurement (assumed to be equal for both groups), *z_crit_* is the critical value for 5% significance =1.96; *z_pwr_* is the power constant, Δ is the relative difference of the means of the variable measured in the two groups^37^. We calculated *z_pwr_* as the inverse of the normal cumulative function in Excel, assuming μ = 0 and σ = 1.

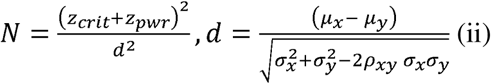

Where N is the total number of pairs and *d* is effect size (Cohen’s *d*). Cohen’s *d* is calculated from μ*_x_* and μ*_y_*, which are means of each visit, σ*_x_* and σ*_y_* are the standard deviations of the two visits, and ρ*_xy_* is the Pearson’s correlation between visits 1 and 2. This is equal to the standard deviation of the differences between the two visits^36,38^.

## Results

In this study we assessed four different datasets spanning healthy controls and PwPs across different time intervals between scans to assess both inter-individual and intra-individual variability in ^31^P-MRS. All spectral data was assessed by the same rater with four years of experience working with MR spectroscopy. The first two datasets were designed to assess short-term variability as well as the effect of MR head coil repositioning. In dataset 1, we conducted three repeated, immediately sequential acquisitions in a neurologically healthy male participant in their 40s without repositioning the subject between scans. In a second single-subject scenario, a neurologically healthy male participant in their 40s was scanned nine times over 24 hours, with repositioning between each session (dataset 2). Datasets three and four assessed the long-term variability in both neurologically healthy controls and PwPs. In dataset 3, five neurologically healthy individuals (3M/2F age 44.6 ± 1.5 years) were scanned at three timepoints: baseline (week 0), week 4, and week 49. Dataset 4 comprised 14 PwPs (13M/1F age 63.7 ± 11.0 years) receiving placebo in the NADPARK study^12^, who underwent scans at baseline and after approximately 30 days (dataset 4). A detailed overview of the datasets is provided in the methods section and illustrated by Figure 1.

Initially, to evaluate the overall variability of cerebral ^31^P-MRS, we assessed the measured metabolite ratios across all four datasets (Figure 2). There were notable inter-individual baseline differences in metabolite ratios across the four datasets, as indicated by the distributions in Figure 2b. Furthermore, intra-individual variability was metabolite-specific, with a clear distinction in distribution and spread between both well-resolved and overlapping metabolites (see methods, Figure 2c). Ratios of well-resolved metabolite signals such as the individual ATP resonances (α, β, γ) and PCr, as well as combined ATP and HEP demonstrated consistently lower intra-individual coefficients of variation (CoV (< 10 %), indicating higher reproducibility (Table 1). In contrast, overlapping metabolites, including PC, GPC, and GPE, showed markedly higher intra-individual variability across the four data sets, with CoVs often exceeding 20 % (Table 1). Notably, the time interval between scans did not exert a consistent influence on variability, as illustrated by the uniform CoV distribution (heatmap in Figure 3). Table 1 provides a full overview of metabolite-specific CoVs across datasets, which are referenced throughout the results.

**Figure 2.**
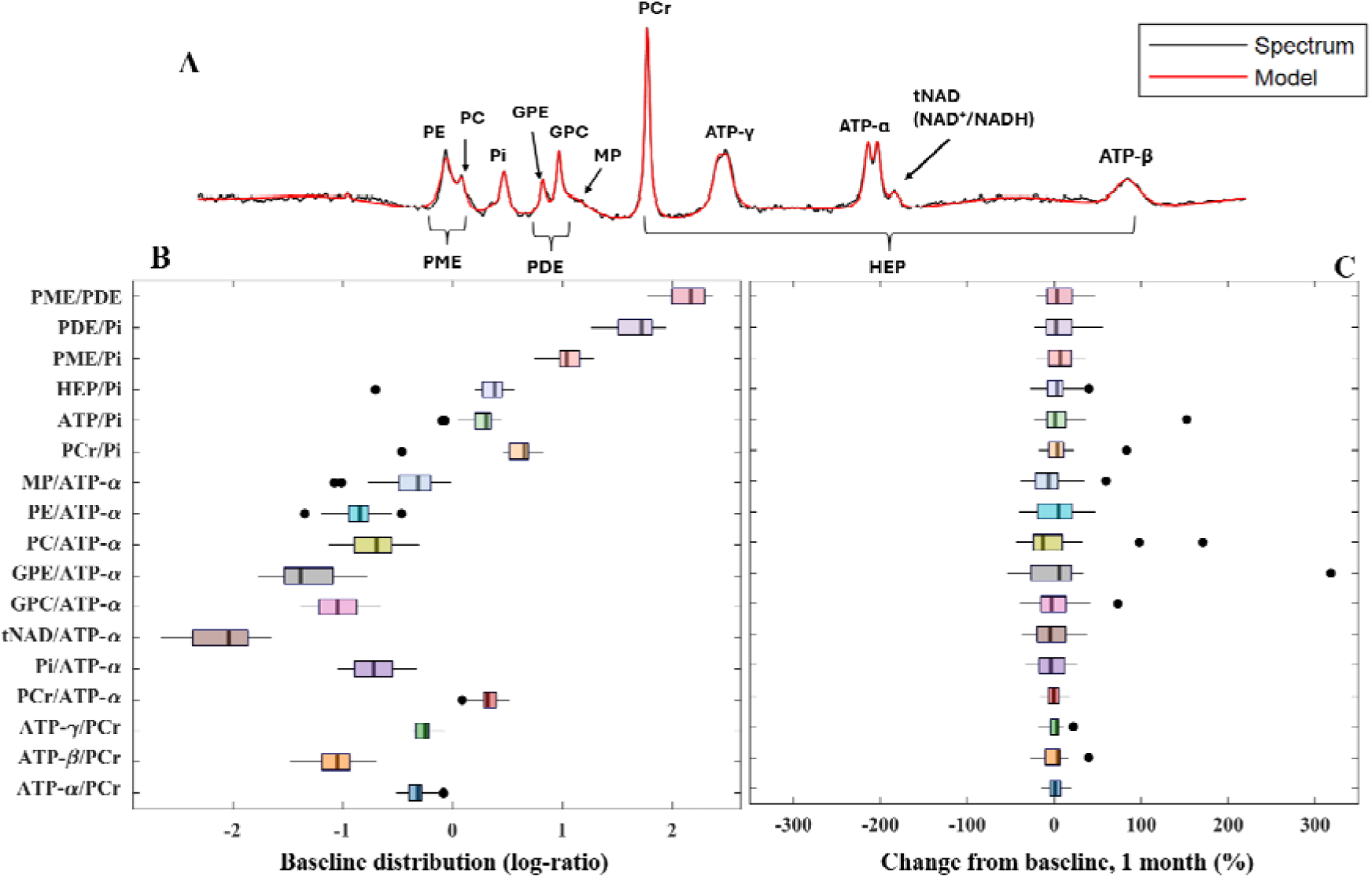
Distribution of ^31^P-MRS measurements across the four datasets. **A:** Example of of a fitted ^31^P-MR spectrum with metabolite signal annotations. **B & C:** Box plots illustrating the distribution and variability of 17 ^31^P-MRS–derived biomarkers measured in the OTP region. **B** shows inter-individual variability (comparison of individual baselines across the four datasets). **C** shows intra-individual variability, measured as percentage change from baseline after 1 month in datasets three and four. The data include healthy participants and persons with Parkinson’s disease, PwPs (n = 21, nPwPs = 14, 18M/3F; mean age 57.3 ± 12.9 years) across four distinct test–retest scenarios. The logarithmic transformation in **C** normalizes skewed ratio distributions and improves visual comparability across metabolites. A log-ratio of zero corresponds to a raw metabolite ratio of 1. A wider box reflects higher variability for any given metabolite ratio. Outliers were defined as values exceeding ±1.5 times the interquartile range (IQR) and are indicated by black dots.

**Figure 3.**
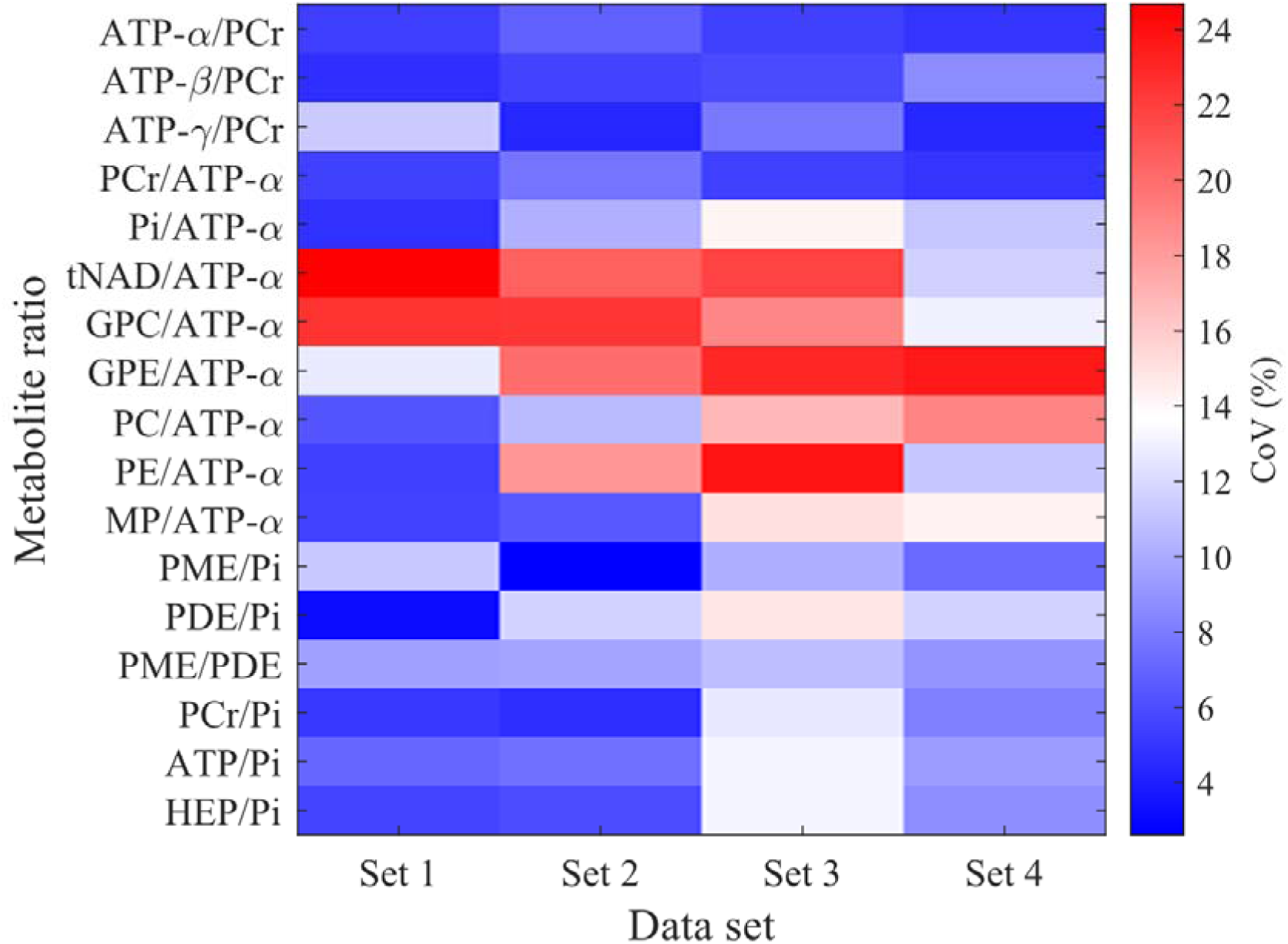
Heatmap of intraindividual test–retest coefficients of variation (CoV%) for 17 metabolite ratios across four data sets. Lower CoV% values (blue) indicate higher test–retest reproducibility; higher values (red) indicate greater variability. For data sets 3 and 4, CoV% was calculated per subject and averaged; for sets 1 and 2, a single CoV% was calculated based on all measurements made in respective sessions.

**Table 1.**
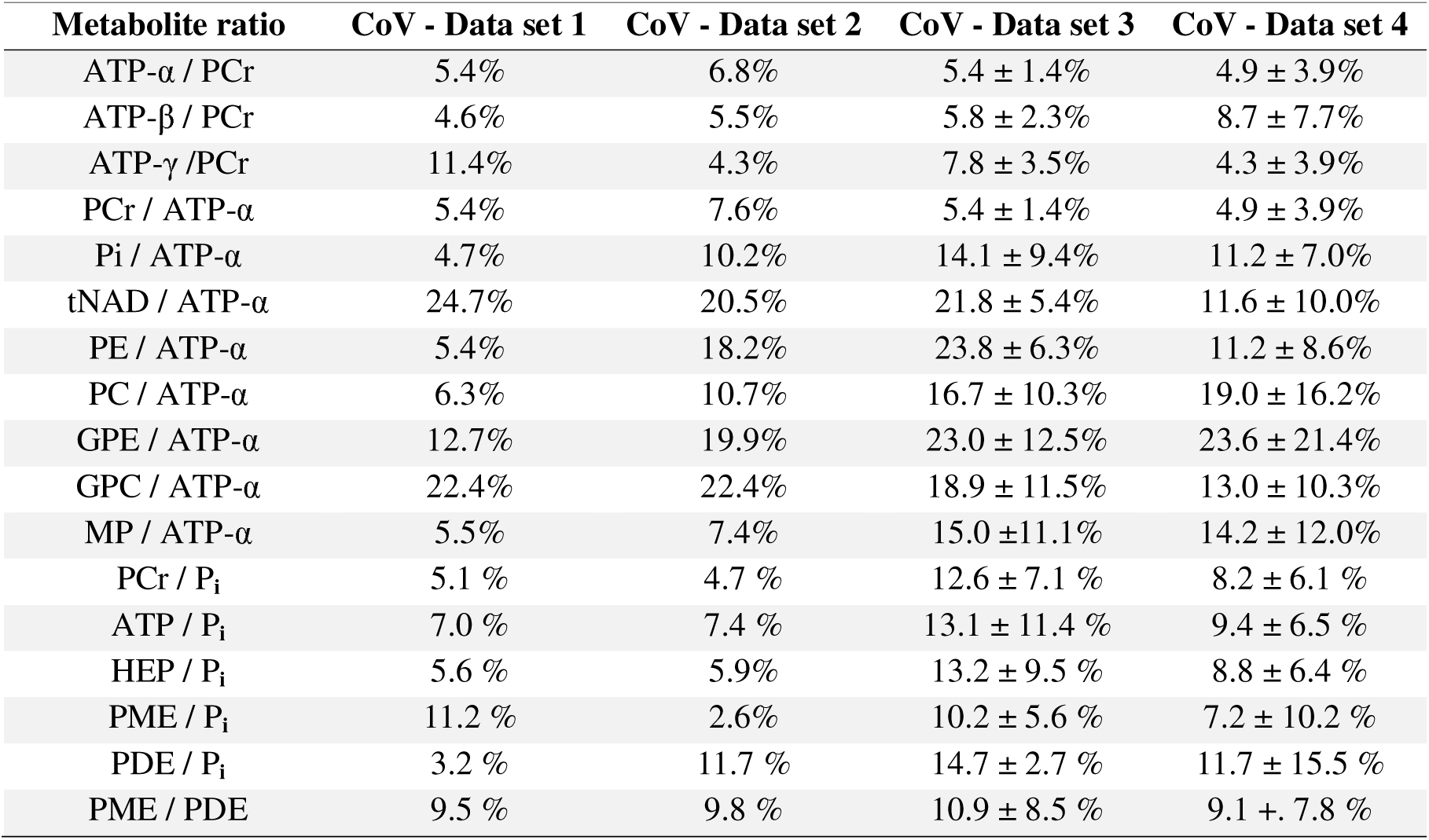
Intra-individual coefficients of variation (CoV%) for metabolite ratios measured across four data sets. CoV% quantifies test–retest variability relative to the mean of each metabolite ratio. For data sets three (n = 5) and four (n = 14), the mean ± SD of individual intra-individual CoV% is reported. Lower values indicate higher stability of the metabolite ratio across test–retest sessions.

| Metabolite ratio | CoV - Data set 1 | CoV - Data set 2 | CoV - Data set 3 | CoV - Data set 4 |
| --- | --- | --- | --- | --- |
| ATP-α / PCr | 5.4% | 6.8% | 5.4 ± 1.4% | 4.9 ± 3.9% |
| ATP-β / PCr | 4.6% | 5.5% | 5.8 ± 2.3% | 8.7 ± 7.7% |
| ATP-γ / PCr | 11.4% | 4.3% | 7.8 ± 3.5% | 4.3 ± 3.9% |
| PCr / ATP-α | 5.4% | 7.6% | 5.4 ± 1.4% | 4.9 ± 3.9% |
| Pi / ATP-α | 4.7% | 10.2% | 14.1 ± 9.4% | 11.2 ± 7.0% |
| tNAD / ATP-α | 24.7% | 20.5% | 21.8 ± 5.4% | 11.6 ± 10.0% |
| PE / ATP-α | 5.4% | 18.2% | 23.8 ± 6.3% | 11.2 ± 8.6% |
| PC / ATP-α | 6.3% | 10.7% | 16.7 ± 10.3% | 19.0 ± 16.2% |
| GPE / ATP-α | 12.7% | 19.9% | 23.0 ± 12.5% | 23.6 ± 21.4% |
| GPC / ATP-α | 22.4% | 22.4% | 18.9 ± 11.5% | 13.0 ± 10.3% |
| MP / ATP-α | 5.5% | 7.4% | 15.0 ± 11.1% | 14.2 ± 12.0% |
| PCr / P<sub>i</sub> | 5.1 % | 4.7 % | 12.6 ± 7.1 % | 8.2 ± 6.1 % |
| ATP / P<sub>i</sub> | 7.0 % | 7.4 % | 13.1 ± 11.4 % | 9.4 ± 6.5 % |
| HEP / P<sub>i</sub> | 5.6 % | 5.9% | 13.2 ± 9.5 % | 8.8 ± 6.4 % |
| PME / P<sub>i</sub> | 11.2 % | 2.6% | 10.2 ± 5.6 % | 7.2 ± 10.2 % |
| PDE / P<sub>i</sub> | 3.2 % | 11.7 % | 14.7 ± 2.7 % | 11.7 ± 15.5 % |
| PME / PDE | 9.5 % | 9.8 % | 10.9 ± 8.5 % | 9.1 ± 7.8 % |

### Intrasession Variability

To assess variability under optimal conditions, we employed a design minimizing physiological and technical variability (dataset 1). Most ratios from well-resolved signals, including ATP/P_i_, HEP/P_i_, ATP-α/PCr, and PCr/P_i_, showed low variability (CoVs <10%), reflecting the robustness of these signals under ideal conditions. Moderate variability (CoVs 10–19%) was observed for ATP-γ/PCr and GPE/ATP-α, while tNAD/ATP-α and GPC/ATP-α exhibited high variability (CoVs 20–29%), consistent with their higher degree of spectral overlap (Table 1).

We then proceeded to introduce potential variation from both coil placement and biological factors (dataset 2). Despite this, ratios from well-resolved signals such as ATP/P_i_, HEP/P_i_, PME/P_i_, and ATP-α/PCr maintained low variability, suggesting resilience to moderate procedural perturbations. Conversely, metabolite ratios from overlapping signals (e.g., tNAD/ATP-α, GPE/ATP-α) again demonstrated higher variability compared to – and consistent with – findings in dataset 1 (Table 1, Figure 3).

### Intersession variability

To assess long-term measurement stability of ^31^P-MRS, we employed dataset 3. Well-resolved metabolite ratios (e.g., ATP-α/PCr, ATP-β/PCr, ATP-γ/PCr) again showed low CoVs, indicating high reproducibility. Moderate variability was observed for P_i_/ATP-α, PC/ATP-α and GPC/ATP-α, while PE/ATP-α, GPE/ATP-α and tNAD / ATP-α exhibited high variability, reinforcing their sensitivity to scan conditions or biological fluctuations (Table 1, Figure 3).

To reproduce this in a neurological disease context, we assessed dataset 4, The variability pattern in this dataset closely mirrored that observed in healthy individuals: well-resolved-metabolite ratios remained highly stable, with low CoVs, whereas overlapping metabolite ratios showed greater intraindividual variability. The tNAD/ATP-α CoV was moderate in this dataset compared to datasets 1-3. Notably, despite differences in disease status, time interval between scanning sessions and demographic composition, the overall variability pattern remained consistent (Table 1, Figure 3). We also found that test-retest reproducibility varied substantially between participants in both datasets 3 and 4, as indicated by the standard deviations of calculated CoVs in table 1.

### Intraclass correlation coefficients (ICCs) and other metrics of test-retest agreement

As complementary measures to CoVs, we examined group-level consistency in datasets 3 and 4 using ICCs, two-groups *t*-tests and Pearson’s correlations. Well-resolved metabolite ratios, as well as tNAD/ATP-α, featured moderate-to-high ICCs (> 0.75) in both datasets, suggesting that a substantial proportion of the group-level variance observed for these measures is attributable to inter-individual differences. The metabolite ratios remained stable between visits across both data sets. An exception was tNAD/ATP-α in dataset 3 which showed a statistically significant difference between weeks 0 and 49 (p = 0.03). Pearson’s correlation coefficients ranged from weak to very strong (e.g., r = 0.03 - 1.0), reflecting heterogeneity in test-retest agreement across metabolites. Full ICC, *t*-test and correlation values are provided in Appendix 1.

To further explore potential systematic bias, we performed a Bland–Altman analysis of dataset 4, which contained the highest number of subjects and best reflected a clinical trial setting. The results were consistent with the CoVs and ICC-based findings. Positive or negative biases were observed for several metabolite ratios including PCr/P_i_, ATP-γ/PCr, ATP/P_i_, tNAD/ATP-α, GPE/ATP-α and PC/ATP-α. For the phosphoester ratios, variability was mainly driven by a single outlying measurement in the second visit of one participant. As no technical error was identified, this data was retained in our variability calculations, but we decided to exclude it from our sample size calculations for matched pairs t-test. In contrast, aggregated metrics such as PME/PDE and HEP/P_i_ showed weak or no bias, indicating high test–retest agreement for these signals. Test-retest bias is illustrated as z-scored differences in figure 4 and as Bland-Altman plots for each metabolite of interest in Appendix 1.

**Figure 4.**
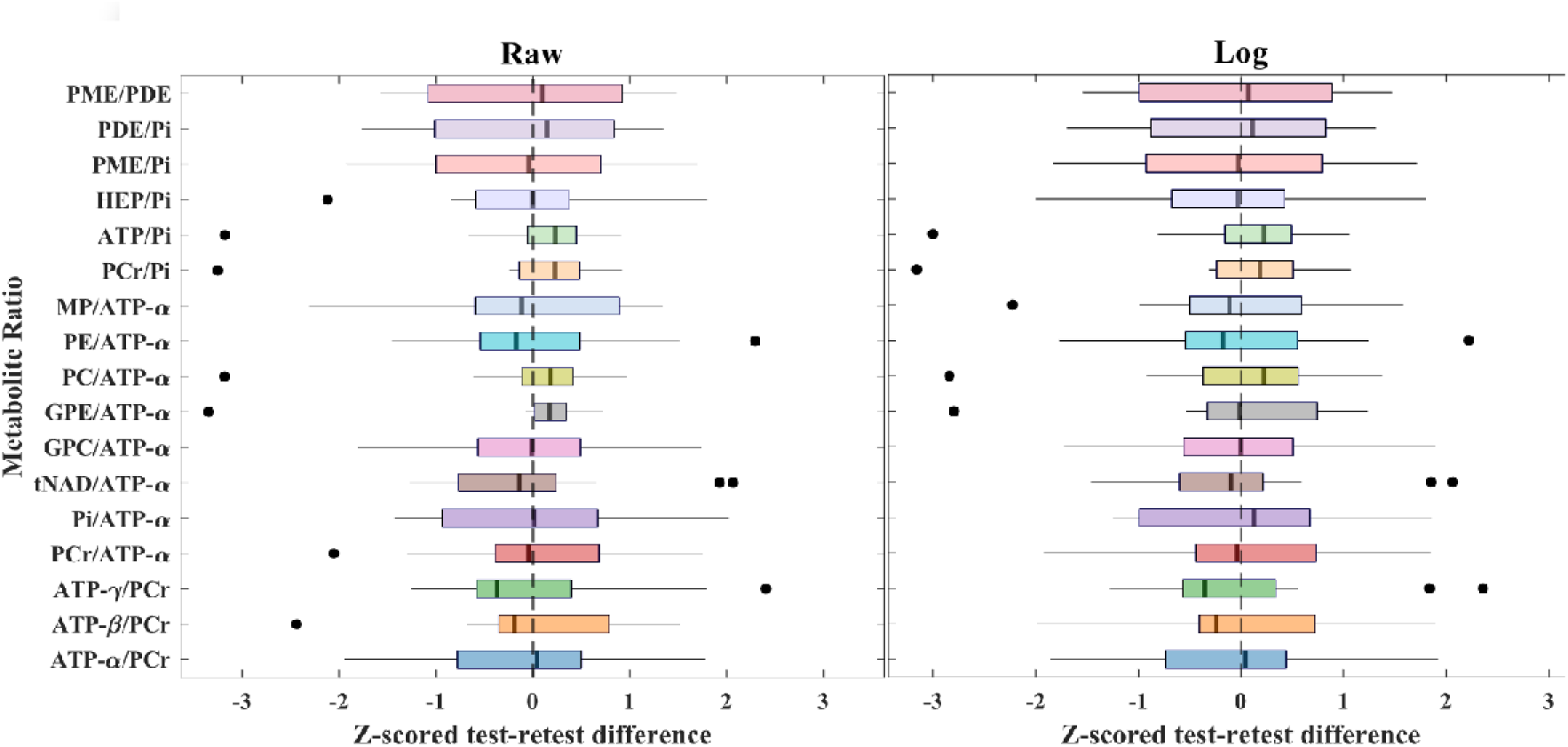
Bland–Altman analysis of metabolite ratio test–retest agreement in data set four. Z-scored intra-individual differences across 17 metabolite ratios, computed from raw (left) and log-transformed (right) values are shown. A box centred around zero indicates minimal test–retest bias, while wider distributions reflect greater variability. Outliers were defined as test–retest differences exceeding ±1.5 times the interquartile range (IQR) and are indicated by black dots. These analyses support the overall repeatability of most metabolite ratios, while highlighting those with greater proportional variability.

To evaluate whether spatial alignment contributed to the observed variability, we assessed voxel-grid placement across repeated scans in the four datasets. Dice similarity coefficients (DSCs) were computed between registered voxel masks for each subject. Mean DSCs exceeded 0.9 in all datasets except dataset 2 (DSC = 0.79), indicating high spatial overlap across measurements. These results suggest that repositioning or registration errors contributed minimally to variability in three of the datasets, while potentially exerting a greater influence in dataset 2. Overall, the findings support the interpretation that observed metabolic variability primarily reflects biological and acquisition-related factors rather than voxel misalignment. DSCs for each visit is provided in Appendix 1.

### Spectral Quality Metrics

To evaluate spectral fitting uncertainty, we calculated the relative Cramér-Rao Lower Bounds (CRLBs) of each fitted metabolite signal amplitude. CRLBs were consistently low (<5%) for PCr and the three ATP resonances, indicating reliable signal fitting. In contrast, tNAD displayed the highest CRLBs, particularly in dataset 1, due to its lower SNR and pronounced spectral overlap with ATP-α. Across datasets, spectral quality tended to improve with larger sample sizes, supporting the utility of group-averaged stability assessments. A complete overview of CRLB values is presented in Appendix 1 – Table 5.

### Sample Size Recommendations

We performed sample size calculations for different study designs using ^31^P-MRS to facilitate study planning. For cross-sectional comparisons, we estimated sample sizes using two-tailed independent-samples t-tests (equal variance), based on baseline data of all participants to capture inter-individual variability. Required sample sizes were calculated for relative changes of 1-100% across statistical power levels of 50-95%. Well-resolved metabolite ratios such as ATP-γ/PCr and ATP-α/PCr yielded the lowest sample-size estimates, supporting feasibility in small cohorts. In contrast, ratios involving overlapping resonances (e.g., tNAD/ATP-α, GPE/ATP-α and PC/ATP-α) required markedly larger cohorts (e.g., n ≥ 27 per group for detecting a 20% change in tNAD/ATP-α), reflecting higher variability.

For longitudinal designs, we used repeated measures from datasets 3 and 4 and applied two-tailed paired samples *t*-tests. Only the first two time-points in dataset 3 were used to ensure comparability with dataset 4. As expected, intra-individual designs reduced sample size requirements relative to cross-sectional designs. Well-resolved metabolite ratios again performed best, while overlapping and aggregate ratios (e.g., GPE/ATP-α, HEP/Pi, PME/PDE) required larger samples, indicating greater intra-individual variability in these measures. Example calculations for 80% power and relative effect sizes in the clinically relevant range (here defined as 10-30%) are detailed in Table 2 and visualized in Figure 5, while the complete set of estimates across all effect sizes and power levels for both *t*-tests is provided in Appendix 3 and 4.

**Figure 5.**
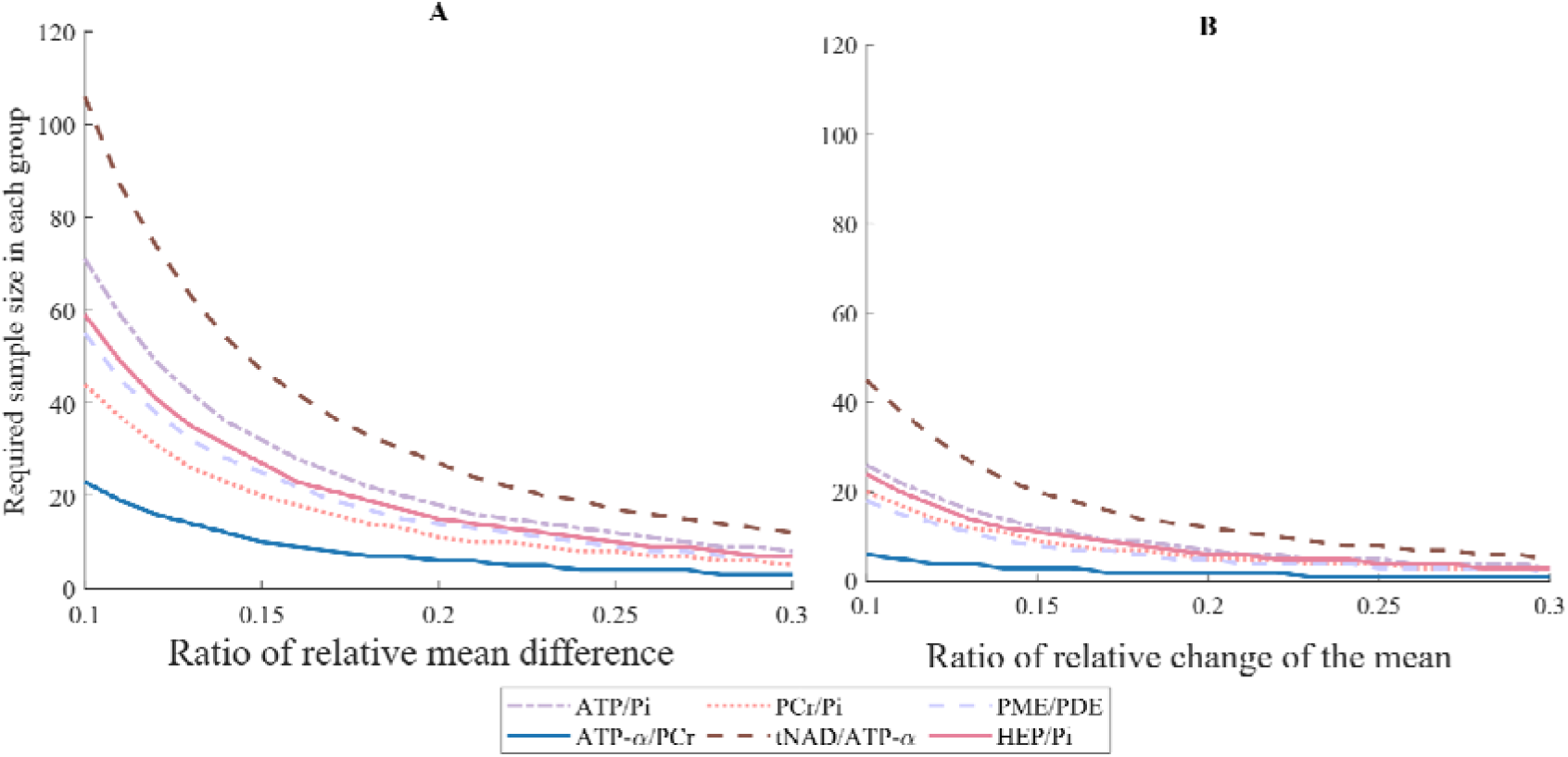
Sample size estimations for commonly assessed metabolite ratios of interest. (A) shows sample sizes required in an independent samples t-test, while (B) shows sample size requirements for a matched pairs t-test. Both calculations assume a statistical power of 80% and a significance level of 0.05. Effects are expressed as ratios (0 to 1) relative to their respective mean difference between groups / change between visits.

**Table 2.** Estimated metabolite-specific required sample sizes for clinically relevant differences. Assuming a statistical power of 80 % and a significance level of 5 %, we present the minimum required number of subjects to detect statistically significant differences in metabolite ratios at mean differences of 10%, 15%, 20%, and 30%. Sample sizes are derived from the observed intra-individual variability in this study. Effect sizes are expressed as percentages of each ratio’s group mean. Lower sample sizes indicate more stable ratios with less intra-individual variability.

| *t-test: Difference between two independent means* |  |  |  |  |  | *t-test: Difference between two dependent means (matched pairs)* |  |  |  |  |
| --- | --- | --- | --- | --- | --- | --- | --- | --- | --- | --- |
| Metab. ratio | Total required sample size N for mean difference |  |  |  |  | Total required sample size N for mean difference |  |  |  |  |
|  | 10% | 15% | 20% | 25% | 30% | 10% | 15% | 20% | 25% | 30% |
| ATP-α/PCr | 46 | 10 | 12 | 8 | 6 | 6 | 3 | 2 | 1 | 1 |
| ATP-β/PCr | 156 | 66 | 38 | 24 | 18 | 17 | 8 | 5 | 3 | 2 |
| ATP-γ/PCr | 26 | 12 | 8 | 6 | 4 | 10 | 5 | 3 | 2 | 2 |
| PCr/ATP-α | 38 | 18 | 10 | 6 | 6 | 6 | 3 | 2 | 1 | 1 |
| Pi/ATP-α | 188 | 84 | 48 | 32 | 22 | 36 | 16 | 9 | 6 | 4 |
| tNAD/ATP-α | 212 | 94 | 54 | 34 | 24 | 45 | 20 | 12 | 8 | 5 |
| PE/ATP-α | 140 | 64 | 36 | 24 | 16 | 58 | 26 | 15 | 10 | 7 |
| PC/ATP-α | 200 | 90 | 50 | 32 | 24 | 65 | 29 | 17 | 11 | 8 |
| GPE/ATP-α | 336 | 150 | 84 | 54 | 38 | 89 | 40 | 23 | 15 | 10 |
| GPC/ATP-α | 178 | 80 | 46 | 30 | 20 | 48 | 22 | 12 | 8 | 6 |
| MP/ATP-α | 166 | 74 | 42 | 28 | 20 | 41 | 18 | 11 | 7 | 5 |
| PME/Pi | 106 | 48 | 28 | 18 | 12 | 8 | 4 | 2 | 2 | 1 |
| PDE/Pi | 72 | 32 | 18 | 12 | 8 | 21 | 9 | 6 | 4 | 3 |
| PME/PDE | 110 | 60 | 28 | 18 | 14 | 18 | 8 | 5 | 3 | 2 |
| PCr/Pi | 88 | 40 | 22 | 16 | 10 | 20 | 9 | 5 | 4 | 3 |
| ATP/Pi | 142 | 64 | 36 | 24 | 16 | 26 | 12 | 7 | 5 | 3 |
| HEP/Pi | 118 | 54 | 30 | 20 | 14 | 24 | 11 | 6 | 4 | 3 |

## 4. Discussion

This study provides a systematic assessment of the test–retest reproducibility of cerebral ^31^P-MRS across four complementary datasets, spanning ideal scanning conditions, longitudinal follow-up in healthy adults, and a RCT cohort of PwPs. Variability was highly metabolite-specific and greater between subjects than within subjects. Ratios derived from well-resolved signals, such as ATP-α, ATP-β, ATP-γ and PCr, consistently showed low coefficients of variation (CoV < 10%) across all settings. In contrast, overlapping metabolite signal ratios (e.g., GPE/ATP-α, GPC/ATP-α, PC/ATP-α and tNAD/ATP-α) exhibited substantially greater variability, often exceeding 20%, irrespective of scan interval, population, or experimental context. These findings suggest that spectral quality, particularly SNR and degree of spectral overlap, rather than study design or participant characteristics, is the principal limitation to reliable quantification. Importantly, our findings are in line with previously reported reproducibility studies featuring different post-processing strategies, anatomical regions of interest, and magnetic field strengths^9,16–18^.

Intrasession reproducibility was generally higher under optimal conditions without repositioning (dataset 1), although some ratios of overlapping metabolites (e.g., tNAD/ATP-α, GPC/ATP-α) still displayed substantial variability in this setting. Conversely, well-resolved metabolite ratios (e.g., ATP-γ/PCr) were in fact more stable in dataset 2, despite repeated repositioning. This underlines that reproducibility is not determined solely by scanning conditions but is strongly metabolite-specific, reflecting the interplay of factors such as spectral overlap and SNR. Lower voxel consistency in dataset 2 (DSC = 0.79 vs. 1.00 in dataset 1) nevertheless supports the interpretation that coil placement and voxel registration contribute to variability. Overall, while experimental setup can modulate reproducibility, intrinsic spectral properties remain the dominant constraint. We conclude that repositioning and time interval between scans had the highest impact on the variability of PME/P_i_, PDE/P_i_, ATP-γ/PCr, P_i_/ATP-α, PE/ATP-α and GPE/ATP-α (Table 1, Figure 3).

Intersession reproducibility showed a mixed pattern across datasets 3 and 4. Our Bland-Altmann analysis revealed that certain metabolite ratios, particularly PCr/P_i_, ATP/P_i_, ATP-γ/PCr, tNAD/ATP-α, PC/ATP-α and GPE/ATP-α, showed modest but consistent bias (Figure 4, Figure S1). Dataset 3, which included a long follow-up interval of ∼45 weeks, was associated with higher variability in several ratios compared to dataset 4, yet this effect was not uniform. This suggests that extended intervals may exacerbate variability, but the magnitude and direction of this effect remain metabolite-dependent. The tNAD/ATP-α ratio differed significantly between weeks 0 and 49 in dataset 3. Importantly, reproducibility patterns in PwPs (dataset 4) were broadly comparable to those observed in healthy participants, indicating that disease status did not exert a consistent additional influence.

Our sample size estimates provide pragmatic benchmarks for future study design. The metabolite-specific variability observed in our reproducibility analyses (Table 1, Figure 3) was reflected in sample size calculations (Table 2, Figure 5). For studies assessing longitudinal effects (e.g.., symptom progression in PD), approximately 20-25 participants are needed to achieve sufficient statistical power for detecting a 20% change. Cross-sectional studies comparing two groups require substantially larger cohorts, highlighting the impact of measurement noise. For example, detecting a between-groups difference of 20% in a moderate-variability metabolite such as tNAD requires approximately 54 participants, which, notably, is higher than the median sample size in prior studies^4^. In both study designs, differences in well-resolved ratios (e.g., ATP-γ/PCr, ATP-α/PCr) could be detected with smaller cohorts, suggesting feasibility for modestly sized studies. In contrast, overlapping metabolite ratios (e.g., GPE/ATP-α, PC/ATP-α, tNAD/ATP-α) required substantially larger cohorts, underscoring the impact of measurement noise until more robust acquisition or quantification methods become available. For studies assessing smaller effect sizes, grouping individual ratios into functional categories (e.g., ATP, HEP, PME, PDE) may improve statistical power at the cost of metabolite-level specificity. These findings highlight the necessity of metabolite-specific power analyses when planning ^31^P-MRS studies.

Several limitations should be acknowledged. First, all acquisitions were performed at a single site on a single scanner with a consistent sequence. While a pre-requisite for our analysis, generalizability may be limited across vendors and field strengths. Sample sizes were modest, particularly for long-term follow-up, though the convergence of variability patterns across datasets supports robustness. Differences in CoV calculation methods (pooled vs. subject-level averages) between single-subject- and multi-subject datasets may also have contributed to discrepancies and should be considered when interpreting results.

Second, dataset 4 contained an unexplained outlier in GPE/ATP-α and PC/ATP-α, which substantially affected the test-retest variability for these metabolite ratios While we found no immediate technical explanation for this outlier, we decided to remove this scan from our sample size calculations to provide more robust estimates. We cannot, however, rule out that there is a biological explanation for this outlier, and therefore we included it in our other test-retest reproducibility analyses.

Third, spectral fitting introduced additional sources of variability. Manual fitting may introduce observer bias, and certain metabolite signals (e.g., tNAD, GPE) showed high relative CRLBs, consistent with overlap and poor SNR at 3T despite NOE-enhancement and ^1^H-decoupling. Higher uncertainty in P_i_ may partly reflect overlap with 2,3-DPG at 5.2 ppm, which was not included in the fitting algorithm^6^. Importantly, CRLBs were reported but not used as rejection criteria to avoid bias, in line with recent recommendations^39^.

Finally, metabolite normalization choices may have influenced results. We normalized our measurements to PCr and ATP-α, due to their low CoVs and CRLBs, in contrast to other studies that normalized to P ^4,10^. While this likely improved stability, potential PCr variability^40^ and ATP-α overlap with tNAD remain potential sources of measurement instability. We found that normalizing to P_i_, as opposed to PCr and ATP-α, introduced higher variability in our datasets. However, to allow for comparison with other studies featuring ^31^P-MRS we calculated and provide metabolite ratios to P_i_ commonly found in literature (e.g., PCr/P_i_). The inclusion of raw metabolite data in the supplementary (Appendix 1) allows investigators to calculate any ratios relevant to their study.

Together, these factors suggest that our sample size estimates and reproducibility measures should be interpreted as indicative benchmarks rather than definitive. Future studies at 3T should pursue cross-site harmonization, automated fitting algorithms, and advanced acquisition strategies to further improve reproducibility of metabolite signals featuring spectral overlap and low SNR.

In conclusion, this study establishes practical reproducibility thresholds for cerebral 31P-MRS and highlights key metabolite-specific constraints that influence study design and interpretability. By identifying which ratios can be reliably quantified over time and under clinical trial conditions, we provide a foundation for standardizing 31P-MRS metrics in both mechanistic and interventional neuroscience research. both research and emerging clinical contexts.

## Acknowledgments

We are grateful to all the participants and staff involved in the study. A special thanks to Max Korbmacher for helpful discussions regarding the paper. We also thank members of the DECODE-PD and the MMIV for their support and fruitful discussions. This study used data from both the NADPARK trial and the NAD-brain study. NADPARK was supported by grants from The Research Council of Norway (288164), Bergen Research Foundation (BFS2017REK05), and the Western Norway Regional Health Authority (F-11470). NAD-brain was supported by grants from the Research Council of Norway (288164), The KG Jebsen Foundation (SKGJ-MED-023), The Western Norway Regional Health Authorities and Norway’s Parkinson Research Fund. This study was also funded by the Western Norway Regional Health Authority through a PhD grant scholarship to Magnus Svensen. The funders had no role in study design, data collection, data analysis and data interpretation. The authors used OpenAI’s ChatGPT (versions 4o and 5) as well as Microsoft’s Co-pilot to assist with language and MATLAB code for figures. All content was reviewed and verified by the authors.

## Author contributions

**M.S –** Designed research. Performed research. Investigation, formal analysis, writing – original draft, writing – review and editing, visualization, performed experiments.

**F.R** Designed research. Investigation, writing – review and editing.

**C.T** Designed research. Investigation, writing – review and editing

**C.D** Designed research. Investigation. Writing – review and editing.

**J.G.S** Writing – review and editing.

**B.B** Investigation, writing – review and editing.

**H.B** Investigation. Writing – review and editing.

**A.C** Software, Writing – review and editing.

**N.B** Protocol and experimental setup, writing – review and editing.

**E.V.S** Collected data. Writing - review and editing.

**V.S** Collected data. Writing – review and editing.

**A.H** Collected data. Writing - review and editing.

## Statements and declarations

Not applicable.

## Data, Materials and Software availability

Anonymized, processed source data are provided with this paper alongside code used for analysis (https://doi.org/10.5281/zenodo.18131110). The raw data are not publicly available due to containing information that could compromise the privacy of research participants. Approval for data sharing is subject to approval by the author’s local ethics committee and a formal data sharing agreement.

## References

1 Schapira AHV. Mitochondrial dysfunction in neurodegenerative disorders. Biochimica et Biophysica Acta (BBA) - Bioenergetics 1998; 1366: 225–233.

2 Forester BP, Berlow YA, Harper DG, Jensen JE, Lange N, Froimowitz MP et al. Age-related changes in brain energetics and phospholipid metabolism. NMR in Biomedicine 2010; 23: 242–250.

3 Covarrubias AJ, Perrone R, Grozio A, Verdin E. NAD+ metabolism and its roles in cellular processes during ageing. Nat Rev Mol Cell Biol 2021; 22: 119–141.

4 Jing Y, Haeger A, Boumezbeur F, Binkofski F, Reetz K, Romanzetti S. Neuroenergetic alterations in neurodegenerative diseases: A systematic review and meta-analysis of *in vivo* 31P-MRS studies. Ageing Research Reviews 2024; 101: 102488.

5 Santos-Díaz A, Noseworthy MD. Phosphorus magnetic resonance spectroscopy and imaging (31P-MRS/MRSI) as a window to brain and muscle metabolism: A review of the methods. Biomedical Signal Processing and Control 2020; 60: 101967.

6 Deelchand DK, Nguyen TM, Zhu X-H, Mochel F, Henry P-G. Quantification of in vivo 31P NMR Brain Spectra using LCModel. NMR Biomed 2015; 28: 633–641.

7 Ren J, Sherry AD, Malloy CR. 31P-MRS of Healthy Human Brain: ATP synthesis, Metabolite Concentrations, pH, and T1 Relaxation Times. NMR Biomed 2015; 28: 1455–1462.

8 de Graaf RA. In Vivo NMR Spectroscopy: Principles and Techniques. 3rd ed. Wiley, 2019.

9 Gordji-Nejad A, Matusch A, Kleedörfer S, Jayeshkumar Patel H, Drzezga A, Elmenhorst D et al. Single dose creatine improves cognitive performance and induces changes in cerebral high energy phosphates during sleep deprivation. Sci Rep 2024; 14: 4937.

10 Prasuhn J, Göttlich M, Ebeling B, Kourou S, Gerkan F, Bodemann C et al. Assessment of Bioenergetic Deficits in Patients With Parkinson Disease and Progressive Supranuclear Palsy Using 31P-MRSI. Neurology 2022; 99: e2683–e2692.

11 Payne T, Appleby M, Buckley E, van Gelder LMA, Mullish BH, Sassani M et al. A doublelblind, randomized, placebolcontrolled trial of ursodeoxycholic acid (UDCA) in Parkinson’s disease. Movement Disorders 2023.https://eprints.whiterose.ac.uk/199907/ (accessed 21 Aug2023).

12 Brakedal B, Dölle C, Riemer F, Ma Y, Nido GS, Skeie GO et al. The NADPARK study: A randomized phase I trial of nicotinamide riboside supplementation in Parkinson’s disease. Cell Metab 2022; 34: 396–407.e6.

13 Ren J, Dewey RB, Rynders A, Evan J, Evan J, Ligozio S et al. Evidence of brain target engagement in Parkinson’s disease and multiple sclerosis by the investigational nanomedicine, CNM-Au8, in the REPAIR phase 2 clinical trials. J Nanobiotechnology 2023; 21: 478.

14 Friebolin H. Basic one- and two-dimensional NMR spectroscopy (5th *completely rev. and enl. ed., pp.* XXIV, 418). Wiley-VCH.

15 Saleh MG, Edden RAE, Chang L, Ernst T. Motion Correction in Magnetic Resonance Spectroscopy. Magn Reson Med 2020; 84: 2312–2326.

16 Parasoglou P, Osorio RS, Khegai O, Kovbasyuk Z, Miller M, Ho A et al. Phosphorus metabolism in the brain of cognitively normal midlife individuals at risk for Alzheimer’s disease. Neuroimage Rep 2022; 2: 100121.

17 Hu S, Yan S, Xie Y, Zhu H, Ding Y, Li Y et al. Test-retest precision of brain metabolites in healthy participants using 31P-MRS and 1H MEGA-PRESS on a 3T multi-nuclear MRI system. Quant Imaging Med Surg 2025; 15: 2852–2864.

18 Lagemaat MW, van de Bank BL, Sati P, Li S, Maas MC, Scheenen TWJ. Repeatability of 31P MR spectroscopic imaging in the human brain at 7 T with and without the Nuclear Overhauser Effect. NMR Biomed 2016; 29: 256–263.

19 McKiernan E, Su L, O’Brien J. MRS in neurodegenerative dementias, prodromal syndromes and at-risk states: A systematic review of the literature. NMR in Biomedicine 2023; 36: e4896.

20 Schmitz B, Wang X, Barker PB, Pilatus U, Bronzlik P, Dadak M et al. Effects of Aging on the Human Brain: A Proton and Phosphorus MR Spectroscopy Study at 3T. J Neuroimaging 2018; 28: 416–421.

21 Cuenoud B, Ipek Ö, Shevlyakova M, Beaumont M, Cunnane SC, Gruetter R et al. Brain NAD Is Associated With ATP Energy Production and Membrane Phospholipid Turnover in Humans. Front Aging Neurosci 2020; 12. doi:10.3389/fnagi.2020.609517.

22 Longo R, Ricci C, Palma LD, Vidimari R, Giorgini A, den Hollander JA et al. Quantitative 31P MRS of the normal adult human brain. Assessment of interindividual differences and ageing effects. NMR in Biomedicine 1993; 6: 53–57.

23 Kemp GJ, Meyerspeer M, Moser E. Absolute quantification of phosphorus metabolite concentrations in human muscle in vivo by 31P MRS: a quantitative review. NMR Biomed 2007; 20: 555–565.

24 Oláh J, Klivényi P, Gardián G, Vécsei L, Orosz F, Kovacs GG et al. Increased glucose metabolism and ATP level in brain tissue of Huntington’s disease transgenic mice. FEBS J 2008; 275: 4740–4755.

25 Moshkova AN, Khvatova EM, Rusakova IA. Analysis and prediction of ATP concentration in the animal brain under hypoxic conditions. Neurochem J 2009; 3: 44–48.

26 Miot F, van Cauteren M, Rooze AK, Geerts A, Osteaux M, Willem R. Non-Invasive in Vivo Determination of the Absolute ATP Concentration in the Rat Liver by 31P NMR Spectroscopy. Bulletin des Sociétés Chimiques Belges 1992; 101: 113–118.

27 Peeters TH, van Uden MJ, Rijpma A, Scheenen TWJ, Heerschap A. 3D 31P MR spectroscopic imaging of the human brain at 3 T with a 31P receive array: An assessment of 1H decoupling, T1 relaxation times, 1H-31P nuclear Overhauser effects and NAD+. NMR in Biomedicine 2021; 34: e4169.

28 Shaka AJ, Keeler J, Freeman R. Evaluation of a new broadband decoupling sequence: WALTZ-16. Journal of Magnetic Resonance (1969) 1983; 53: 313–340.

29 Purvis LAB, Clarke WT, Biasiolli L, Valkovič L, Robson MD, Rodgers CT. OXSA: An open-source magnetic resonance spectroscopy analysis toolbox in MATLAB. PLOS ONE 2017; 12: e0185356.

30 Vanhamme L, van den Boogaart A, Van Huffel S. Improved Method for Accurate and Efficient Quantification of MRS Data with Use of Prior Knowledge. Journal of Magnetic Resonance 1997; 129: 35–43.

31 Edden RAE, Puts NAJ, Harris AD, Barker PB, Evans CJ. Gannet: A batch-processing tool for the quantitative analysis of gamma-aminobutyric acid–edited MR spectroscopy spectra. J Magn Reson Imaging 2014; 40: 1445–1452.

32 Jm B, Dg A. Statistical methods for assessing agreement between two methods of clinical measurement. Lancet (London, England) 1986; 1.https://pubmed.ncbi.nlm.nih.gov/2868172/ (accessed 19 Jun2024).

33 Bland JM, Altman DG. Applying the right statistics: analyses of measurement studies. Ultrasound Obstet Gynecol 2003; 22: 85–93.

34 Pratt J, McStravick J, Kennerley AJ, Sale C. Intra- and inter-session reliability and repeatability of 1H magnetic resonance spectroscopy for determining total creatine concentrations in multiple brain regions. Experimental Physiology; n/a. doi:10.1113/EP092252.

35 McGraw KO, Wong SP. ‘Forming inferences about some intraclass correlations coefficients’: Correction. Psychological Methods 1996; 1: 390–390.

36 (PDF) GPOWER: a general power analysis program. ResearchGate 2024. doi:10.3758/BF03203630.

37 Eng J. Sample Size Estimation: How Many Individuals Should Be Studied? Radiology 2003; 227: 309–313.

38 Layla. Correcting Two-Sample z and t Tests for Correlation: An alternative to One-Sample Tests on Difference Scores. Psicológica. 2012.https://psicologicajournal.com/correcting-two-sample-z-and-t-tests-for-correlation-an-alternative-to-one-sample-tests-on-difference-scores/ (accessed 16 Sep2025).

39 Kreis R. The trouble with quality filtering based on relative Cramér-Rao lower bounds. Magnetic Resonance in Medicine 2016; 75: 15–18.

40 Li BSY, Wang H, Gonen O. Metabolite ratios to assumed stable creatine level may confound the quantification of proton brain MR spectroscopy. Magnetic Resonance Imaging 2003; 21: 923–928.

